# Whole exome sequencing of a cohort of patients with refractory JIA reveals rare genetic variants for paediatric monogenic diseases

**DOI:** 10.64898/2026.01.30.26345195

**Authors:** Melissa Tordoff, Samantha L. Smith, Gillian Rice, Saskia Lawson-Tovey, Nisha Nair, Lianne Kearsley-Fleet, Andrew D Smith, UK JIA Biologics Register, CAPS, CLUSTER, Tracy A Briggs, Athimalaipet V. Ramanan, Andrew P. Morris, Stephen Eyre, Kimme L. Hyrich, Lucy R. Wedderburn, John Bowes, the CLUSTER consortium

## Abstract

**Objectives:** Research of refractory disease in juvenile idiopathic arthritis (JIA) is limited, and a potential genetic contribution has yet to be investigated. This study aimed to explore the presence of rare monogenic disease gene coding variants in a refractory JIA population.

**Methods:** Cases were included with a record of inefficacy for methotrexate and ≥1 biologic drug or exposure to methotrexate and ≥2 biologic drugs for any reason. Whole exome sequencing data were analysed using VarSeq. rarity and pathogenicity filters were applied. Variants within an OMIM curated paediatric monogenic gene list, arthritis OMIM gene list, primary immunodeficiency gene panel (PanelApp) or gene reported for JIA drug response or toxicity (ClinPGX) were retained. ACMG classification excluded benign or likely benign variants.

**Results:** In total, 83 individuals were included. Twelve variants were previously reported in other paediatric onset diseases with similar phenotypes to JIA. Seventeen variants were detected in twelve genes with an arthritis OMIM phenotype. Seventeen variants were detected within fourteen genes that were reported on the primary immunodeficiency panel (PanelApp) and were previously reported in a publication. A total of 39 variants were detected in genes from a JIA drug response or toxicity gene list (ClinPGX).

**Conclusions:** This study evidences that 66 individuals with refractory JIA carry rare variants associated with paediatric diseases, JIA susceptibility loci or drug response and toxicity. These variants could contribute to refractory disease, mimics of JIA/complicated phenotypes or effect treatment response. Longitudinal data are needed to confirm these findings.

## Introduction

Juvenile idiopathic arthritis (JIA) is the most common inflammatory rheumatic disease of childhood. The term refers to a group of heterogenous arthritides, with symptoms that last more than 6 weeks with onset before the age of 16 (1). Unlike adult rheumatoid arthritis (RA), where recent EULAR guidelines define difficult-to-treat, refractory disease does not currently have a standardised definition for JIA. However, one study reported that definitions of refractory disease in RA and JIA often include resistance to multiple drugs with multiple mechanisms of action and persistence of symptoms and high disease activity (2). Previous literature estimated that around 5-10% of individuals with systemic and polyarticular JIA have refractory disease, which can result in severe joint destruction, growth retardation, psychosocial morbidity and adverse effects from long-term use of immunosuppressive drugs (3). A more recent study of treatment and outcomes of JIA reported that 17% of JIA patients’ disease did not achieve minimal disease activity using cJADAS10 criteria within 70 weeks of diagnosis (4). Furthermore, regardless of whether they achieved the outcome, one in five patients in the study had improvement in joint counts and physician assessment of disease despite consistent poor patient reported scores on pain and functional disability (5).

A recent study of biologic drug use in JIA reported that approximately 5% of patients starting their first biologic drug went on to have biologic refractory disease (patients were classified as refractory upon starting a third biologic) (6). Additionally, a study of refractory disease in systemic JIA (sJIA) found that despite advancements in biologic treatments, 20% of sJIA patients have refractory disease (7). Despite this evidence of refractory disease in JIA patients, there is a significant absence of research of refractory disease in childhood rheumatic disease. One study noted that the proportion of publications investigating refractory disease were much higher in RA than in polyarticular JIA (2), which highlights the need for further research of refractory JIA.

Genetic testing of refractory JIA may inform clinicians and researchers of mechanisms behind refractory disease and personalised treatment options. Evidence in case studies demonstrate that misclassification of JIA can present as “difficult to treat” and further diagnostic testing reveals a different paediatric disease of similar phenotype. Coexisting conditions may also contribute to difficult treatment response (see Supplementary Table 1). One case study reports a patient diagnosed with JIA with continued symptoms after prolonged non-steroidal anti-inflammatory usage. Genetic analysis revealed a homozygous mutation of the *WISP3* (*CCN6*) gene leading to a change in diagnosis to progressive psuedorheumatoid dysplasia, which informed physical therapy and treatments. Another case study of a child with refractory polyarticular JIA demonstrates that genetic testing aided a secondary diagnosis of Leri Weil syndrome. This individual had long-term adverse effects of glucocorticoids and was able to access more effective treatments as a result of this diagnosis (9). Monogenic disorders of the PRG4 gene can lead to a severe deforming arthropathy phenotype and have been detected in several case series of cases initially thought to be ‘difficult-to-treat-JIA’(10). Finally, one case study reports a three-generation family of rheumatoid factor positive polyarthritis that, through the use of sequencing, was later diagnosed as STING-associated vasculopathy with onset in infancy (SAVI)- an interferonopathy (11). Therefore, the first aim of this study was to investigate the presence of rare genetic variants in paediatric monogenic disease genes within a cohort of patients with refractory JIA.

A second possible contribution to refractory JIA is the presence of rare genetic variants for JIA that manifest in a subtype unresponsive to current treatments for the disease. Genome-wide association studies (GWAS) in JIA have defined susceptibility through the Human Leukocyte Antigen (HLA) region of the genome and 22 additional loci outside of the *HLA* region (12–14). Moreover, monogenic familial forms of JIA have been reported, linked to the genes *LACC1, LRBA, NFIL3* and *UNC13D* (*15*). Monogenic forms can be characterised by an early onset, familial clustering and increased disease severity, as seen in inflammatory diseases such as systemic lupus erythematosus (SLE) (16). Therefore, the second aim of this study was to investigate the presence of rare genetic variants for JIA in monogenic genes linked to arthritis.

Finally, refractory JIA could be contributed to by genetic variants affecting response or toxicity to common JIA treatments such as methotrexate and TNF inhibitors. Pharmacogenetic studies have long discovered genetic variants associated with drug response. One example being genetic variation in the *ATIC* gene being associated with poor response to methotrexate in JIA (17). Mutations such as these could contribute to refractory disease. Therefore, the final aim of this study was to investigate the presence of rare genetic variants in genes linked to drug response and toxicity of JIA treatment drugs.

## Methods

### Sample selection

Cases were selected from around 4300 JIA samples as part of the CLUSTER consortium. These were recruited to the following UK-wide JIA cohort studies: the UK JIA Biologics Register, which includes the British Society for Paediatric and Adolescent Rheumatology ETN cohort study (BSPAR-ETN) and the Biologics for Children with Rheumatic Diseases study (BCRD) (18) and the Childhood Arthritis Prospective Study (CAPS) (19). Samples were prioritised for sequencing if DNA was available and if one of the following treatment response criteria was met: 1) both methotrexate treatment had failed for inefficacy as well as ≥1 biologic drug had been discontinued with a recorded reason of inefficacy, or 2) if the individual had exposure to methotrexate and exposure to ≥2 biologic drugs for any reason. Records were historical and further biological and clinical data could not be collected. All participants provided written informed consent, or age-appropriate assent with written informed parental consent.

### Sequencing

Whole exome sequencing (WES) was performed using the Agilent SureSelect Human All ExonV6 kit (Agilent, Santa Clara, CA) according to the manufacturer’s instructions by Novogene, UK (Cambridge). Paired-end sequencing, resulting in 150 bases from each end of the DNA fragments was performed using the Novaseq 6000 Genome analyser (Illumina, San Diego, CA). Sequencing had a 100x sequencing coverage. of disease Quality control was conducted by Novogene and included the removal of paired-read ends if either read contained adaptor contamination, if more than 10% of bases were uncertain or if the proportion of low-quality bases was over 50%. Sequencing error rate, quality distribution and GC content distribution were assessed to ensure quality data and the clean data were then aligned to the human reference genome (hg38).

### Variant annotation and filtering

Variant annotation and filtering were performed in VarSeq^TM^ (v2.2.1) (20), Figure 1. Variants with a read depth ≥30 and genotype quality ≥80 were retained for analysis. Rarity and pathogenicity filters were applied using VarSeq to prioritise likely pathogenic variants, method adapted from Belot et al., 2020 (21). Rarity filters were applied following annotation of variants with the allele frequency databases; ExAC variant frequencies 1.0 (22), gnomAD Exomes Variant Frequencies 2.0.1 (23), gnomAD Genomes Variant Frequencies 2.0.1 v2, NHLBI ESP6500SI-V2-SSA137 Exomes Variant Frequencies 0.0.30 (24) and 1kG Phase 3 (25). Variants with an allele frequency <1% or missing in public frequency datasets were retained for analysis. Pathogenicity filters were applied to the variants using the databases ClinVar, CADD v1.5, REVEL functional predictions, RefSeq Genes V1 and dbscSNV splice altering predictions 1.1. Variants classified as benign or likely benign by ClinVar and had a review status greater than three stars were removed from further analysis (26). Variants with a CADD PHRED score ≥15 and a REVEL score ≥0.7 were retained for analysis. Variants classified as loss of function or missense using RefSeq and variants with an Ada score ≥0.6 and RF score ≥0.6 from dbscSNV splice altering predictions were retained for analysis. Only single nucleotide variants were analysed. According to The American College of Medical Genetics and Genomics (ACMG) classification criteria for the interpretation of sequence variants (27), variants can be classified as pathogenic, likely pathogenic, uncertain significance likely benign or benign. Further information on ACMG guidelines can be found in Richards *et al.,* 2015 (27). These guidelines aid in the prioritisation of candidate causative variants, however it is recommended that additional clinical evaluation is required to support diagnoses (27). Variants classified as likely benign or benign for ACMG classification by VarSeq were excluded from the analysis. Variants are presented with allele frequency of Non-Finnish European populations from UKBiobank.

**Figure 1.**
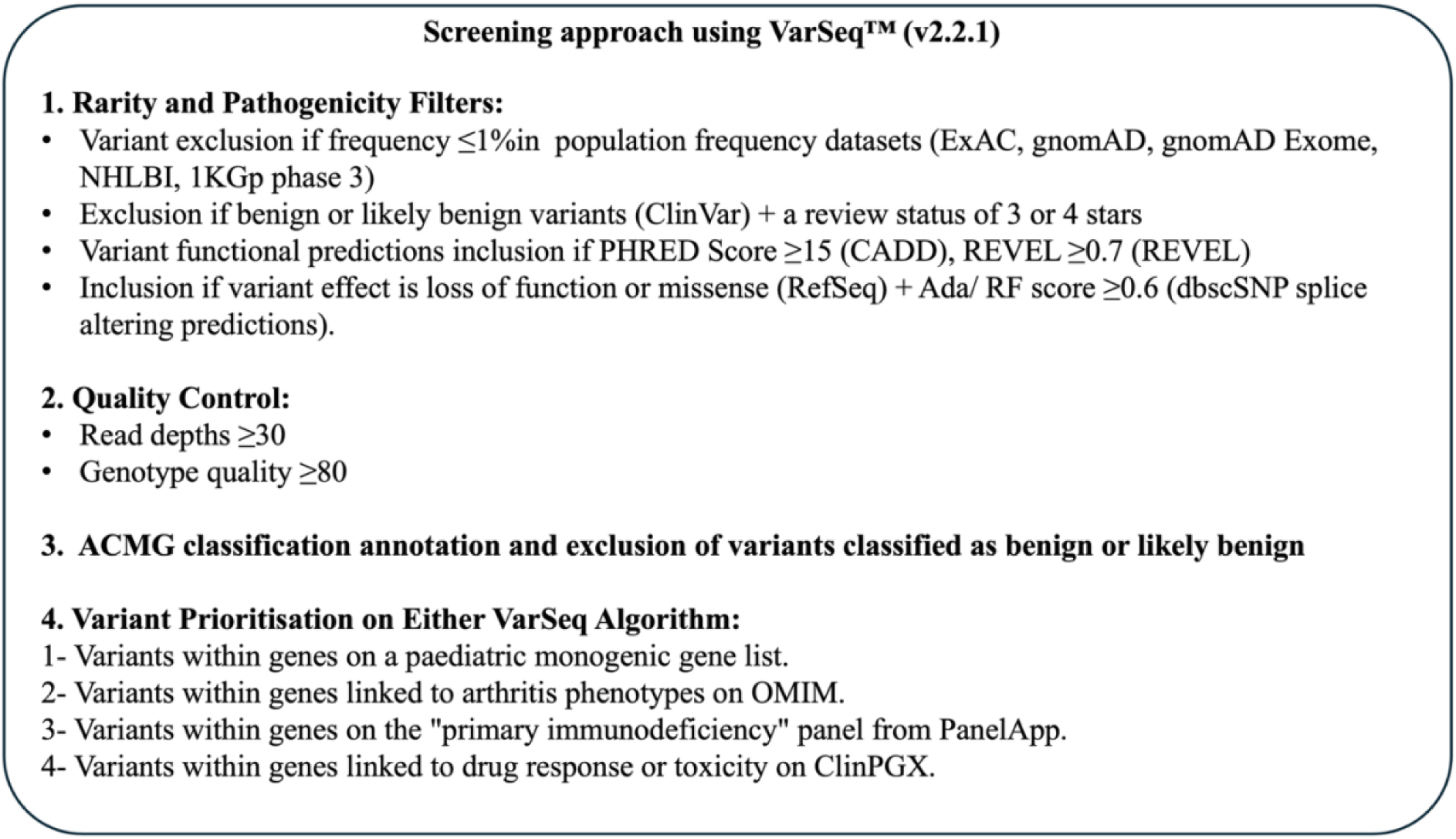
Filtering Pipeline for Prioritising Variants.

### Prioritising Variants

Three VarSeq algorithms were implemented to prioritise variants relevant to the research questions (Figure 1). Algorithm one prioritised variants within genes that appeared in a paediatric monogenic gene list supplied to the VarSeq software. To create a paediatric monogenic gene list, the Medical subject headings (MeSH) database was used to create standardised search terms for juvenile arthritis (28). The MeSH database was searched for juvenile arthritis and the subheadings that juvenile arthritis produced were used as search terms. These search terms were then implemented in the online Mendelian inheritance in man (OMIM) database to generate a list of genes linked to monogenic diseases (29). Algorithm two prioritised variants within genes that listed one of the following phenotypes in the OMIM database: “Arthralgia/Arthritis, Juvenile Rheumatoid Arthritis, Oligoarthritis, Polyarticular Arthritis, Rheumatoid Arthritis” (29). Algorithm three prioritized variants within genes that were present on the “primary immunodeficiency” v2.1 panel from PanelApp (30). Finally, the ClinPGX database (31) was consulted for mutations and genes linked to the drugs abatacept, adalimumab, anakinra, canakinumab, etanercept, golimumab, infliximab, methotrexate, rituximab and tocilizumab, as per the treatment records of the cohort. Variants and genes listed for relevant drug response or toxicity were then prioritized in the final filtered dataset. The total number of unique variants (variants could appear in more than one algorithm) that appeared in at least one algorithm or drug response or toxicity gene was 399 across 287 genes. All remaining variants were then assessed for disease contribution or drug response and/or toxicity using available literature.

## Results

### Study population

A total of 83 individuals fulfilled the inclusion criteria and were selected for WES (Table 1): 50 according to definition 1 and 33 by definition 2. Table 1 displays the proportion of individuals per International League of Associations for Rheumatology (ILAR) subtype and sex per inclusion group for the cohort and the median age at onset (AAO) for JIA in each group. In total, 75 (90%) individuals in this cohort were of white European ancestry. A total of 66 (79%) samples within this cohort carried a variant that passed the filters variant prioritisation pipeline and appeared in one of the four variant prioritisation gene lists/algorithms.

**Table 1.** Population characteristics of 83 individuals with refractory JIA. The number and percentage of individuals in each ILAR subtype, sex and mean age of onset for JIA per inclusion criteria in the cohort. Total number of individuals= 83. Percentage of individuals per the total number of individuals in the population (83). To protect identities, number of individuals <5 is not presented in the table. ILAR; international league of associations for rheumatology, MTX; methotrexate, AAO; age at onset. *1 sample has missing AAO data in the group “MTX (inefficacy) ≥1 biologic (inefficacy)”, 4 samples had missing AAO data in the group “Exposure MTX ≥2 biologic (for any reason)”.

| **ILAR Subtype n (%)** | **MTX (inefficacy)**<br>**≥1 biologic**<br>**(inefficacy)** | **Exposure MTX ≥2**<br>**biologic (for any**<br>**reason)** |
| --- | --- | --- |
| Systemic Arthritis | 6 (7%) | 13 (16%) |
| Oligoarthritis | 16 (19%) | 5 (6%) |
| RF Negative Polyarthritis | 15 (18%) | <5 |
| RF Positive Polyarthritis | 6 (6%) | 0 |
| Psoriatic Arthritis | <5 | 0 |
| Enthesitis Related Arthritis | <5 | <5 |
| Undifferentiated Arthritis | <5 | 10 (12%) |
| **Sex n (%)** |  |  |
| Male | 15 (18%) | 9 (11%) |
| Female | 35 (42%) | 24 (29%) |
| **AAO Median (SD), yrs** | **7.5 (4.9)\*** | **4.4 (4.4)\*** |
| Total n | 50 | 33 |

### Variants previously reported in paediatric diseases

A total of 300 variants were prioritised in algorithm one across 204 genes and 63 patients. ClinVar was used to search for literature associated to each variant and variants with evidence of pathogenicity to a paediatric disease with phenotype similar to JIA were included in Table 2. A total of 12 variants at 8 genes in 11 individuals had been previously reported in a paediatric disease that had an overlapping phenotype to JIA. One individual in the cohort carried two of these variants within *ATP7B*: p.Ille1148Thr and p.Gln1142His. Of the 12 variants, only one variant, p.Arg999His in *RYR1,* was detected in the same genotype that has been reported in the literature. The remaining variants reported in Table 2 were not detected in this JIA cohort in the same genotype reported for the corresponding disease in the literature. The remaining variants prioritised by algorithm one are reported in Supplementary Table 2.

**Table 2.**
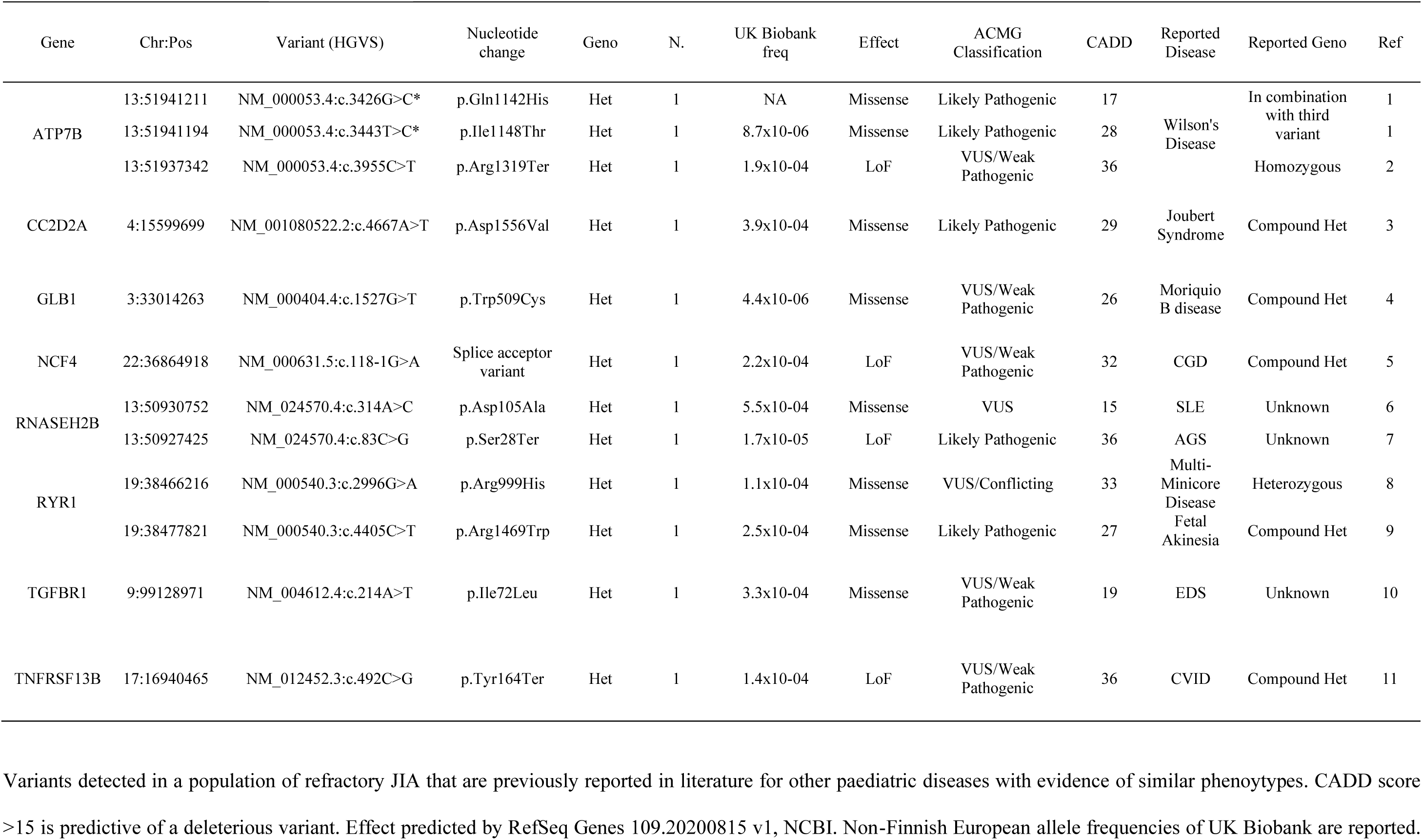
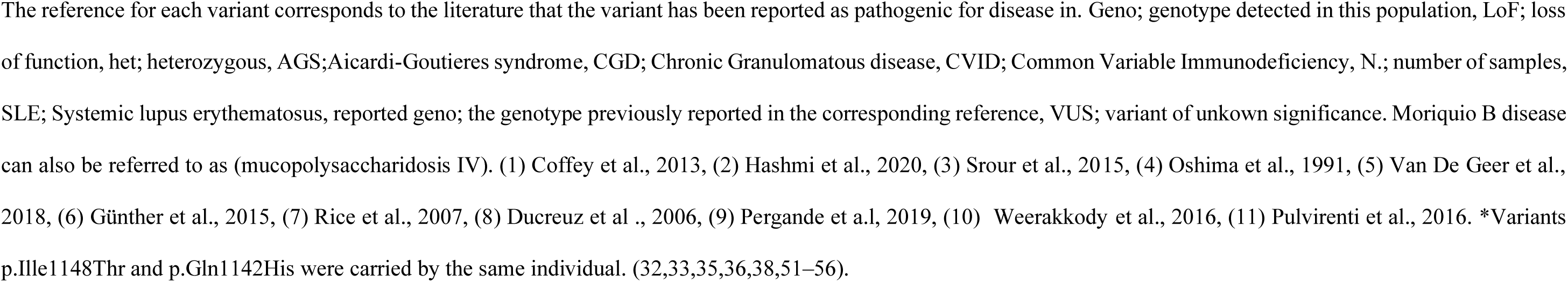
Variants within genes in a paediatric monogenic gene list that have been previously reported and have evidence of misclassification with JIA (algorithm 1). Variants detected in a population of refractory JIA that are previously reported in literature for other paediatric diseases with evidence of similar phenoytypes. CADD score >15 is predictive of a deleterious variant. Effect predicted by RefSeq Genes 109.20200815 v1, NCBI. Non-Finnish European allele frequencies of UK Biobank are reported. The reference for each variant corresponds to the literature that the variant has been reported as pathogenic for disease in. Geno; genotype detected in this population, LoF; loss of function, het; heterozygous, AGS;Aicardi-Goutieres syndrome, CGD; Chronic Granulomatous disease, CVID; Common Variable Immunodeficiency, N.; number of samples, SLE; Systemic lupus erythematosus, reported geno; the genotype previously reported in the corresponding reference, VUS; variant of unkown significance. Moriquio B disease can also be referred to as (mucopolysaccharidosis IV). (1) Coffey et al., 2013, (2) Hashmi et al., 2020, (3) Srour et al., 2015, (4) Oshima et al., 1991, (5) Van De Geer et al., 2018, (6) Günther et al., 2015, (7) Rice et al., 2007, (8) Ducreuz et al., 2006, (9) Pergande et a.l, 2019, (10) Weerakkody et al., 2016, (11) Pulvirenti et al., 2016. *Variants p.Ille1148Thr and p.Gln1142His were carried by the same individual. (32,33,35,36,38,51–56).

### Variants within Genes matched to OMIM phenotypes

A total of 17 variants were prioritised in algorithm two across 12 genes and 17 patients, Table 3. Algorithm two detected variant within genes listed on OMIM with a phenotype matching “Arthralgia/Arthritis, Juvenile Rheumatoid Arthritis, Oligoarthritis, Polyarticular Arthritis or Rheumatoid Arthritis”, Table 3. Four variants in algorithm two have been previously reported based on ClinVar literature. *ATP7B* variants p.Gln1142His, p.Ile1148Thr and p.Arg1319Ter have been previously reported in studies of Wilson’s disease (32,33). *HNF1B* variant p.Met532Val is linked to maturity onset diabetes of the young type 5 (MODY5) (34).

**Table 3.**
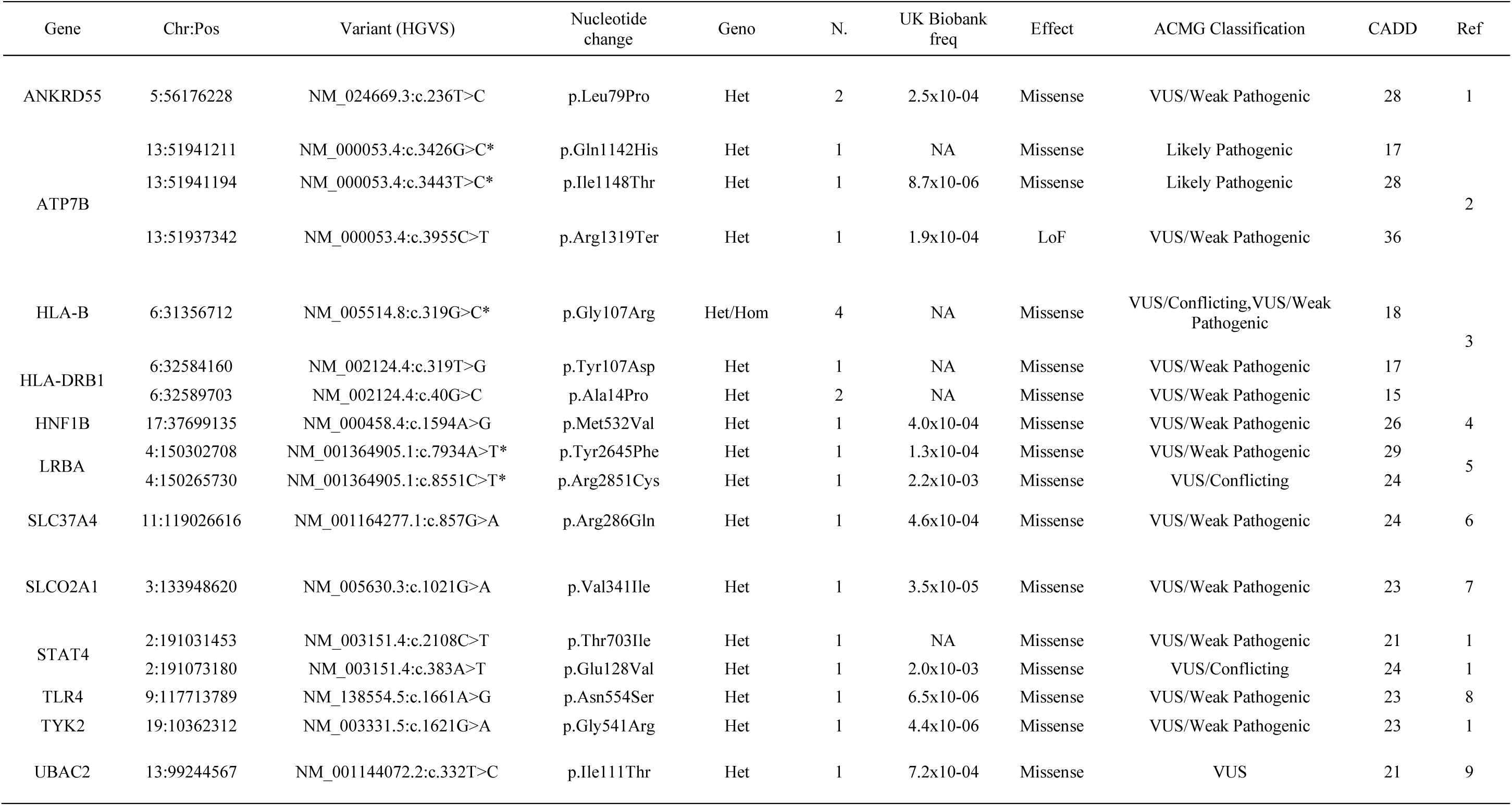
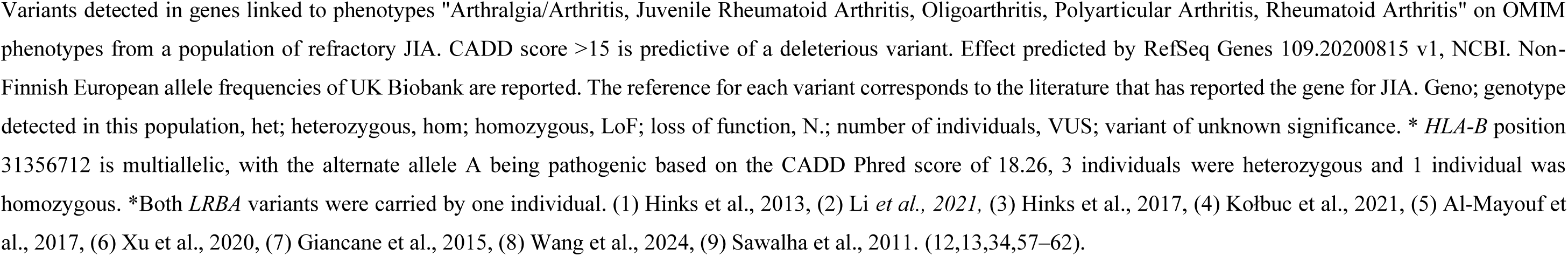
Variants in genes matched to arthritis phenotypes using OMIM (algorithm 2) with evidence of gene pathogenicity to arthritis in literature. Variants detected in genes linked to phenotypes "Arthralgia/Arthritis, Juvenile Rheumatoid Arthritis, Oligoarthritis, Polyarticular Arthritis, Rheumatoid Arthritis" on OMIM phenotypes from a population of refractory JIA. CADD score >15 is predictive of a deleterious variant. Effect predicted by RefSeq Genes 109.20200815 v1, NCBI. Non-Finnish European allele frequencies of UK Biobank are reported. The reference for each variant corresponds to the literature that has reported the gene for JIA. Geno; genotype detected in this population, het; heterozygous, hom; homozygous, LoF; loss of function, N.; number of individuals, VUS; variant of unknown significance. * *HLA-B* position 31356712 is multiallelic, with the alternate allele A being pathogenic based on the CADD Phred score of 18.26, 3 individuals were heterozygous and 1 individual was homozygous. *Both *LRBA* variants were carried by one individual. (1) Hinks et al., 2013, (2) Li *et al., 2021,* (3) Hinks et al., 2017, (4) Kołbuc et al., 2021, (5) Al-Mayouf et al., 2017, (6) Xu et al., 2020, (7) Giancane et al., 2015, (8) Wang et al., 2024, (9) Sawalha et al., 2011. (12,13,34,57–62).

### Variants within “Primary Immunodeficiency” Genes

A total of 121 variants were prioritised in algorithm three across 92 genes and 54 patients. ClinVar was utilised to search the literature for each variant and variants that were previously reported in a disease are presented in Table 4. A total of 15 variants across 14 genes and 16 patients were detected within genes listed on the primary immunodeficiency panel from PanelApp that had been previously reported in the literature. The remaining novel variants that were prioritised in algorithm three are reported in Supplementary Table 2.

**Table 4.**
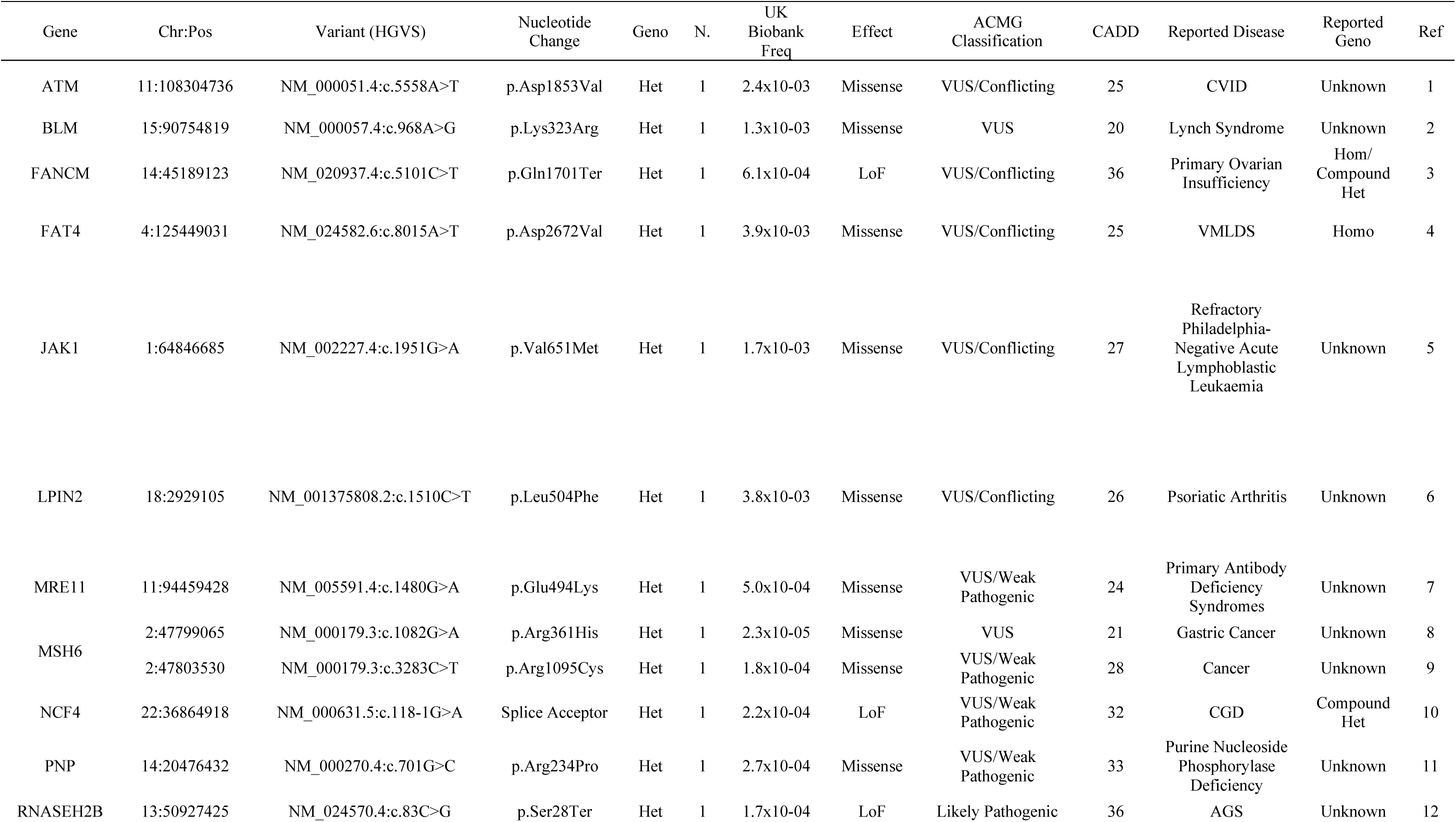
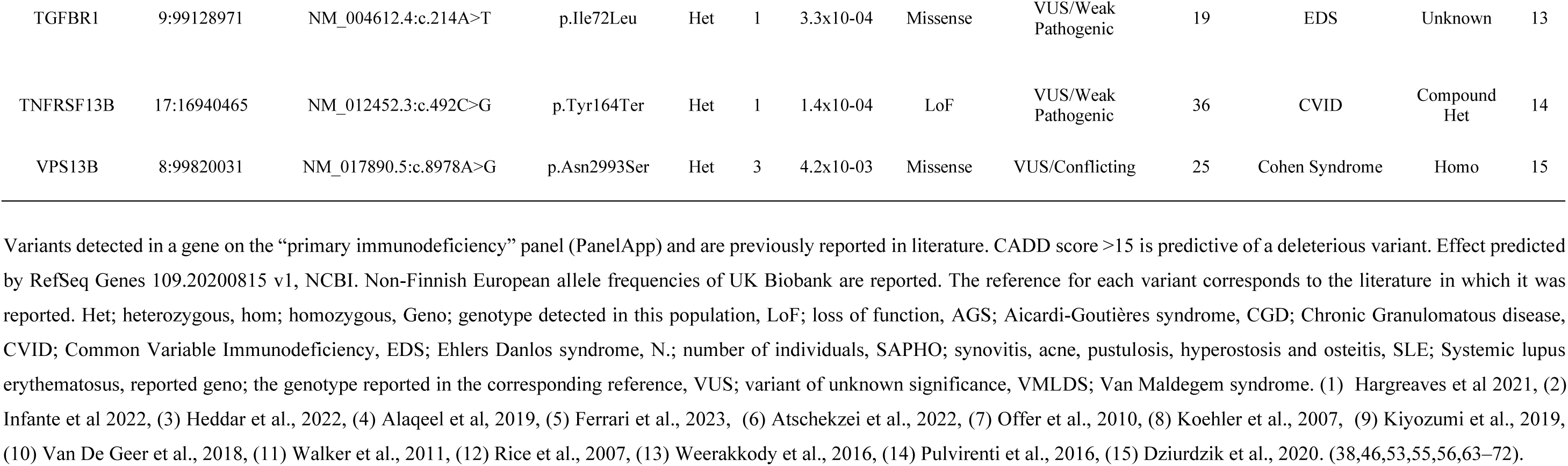
Variants detected within genes on the primary immunodeficiency panel from PanelApp (algorithm 3) that have been previously reported in literature. Variants detected in a gene on the “primary immunodeficiency” panel (PanelApp) and are previously reported in literature. CADD score >15 is predictive of a deleterious variant. Effect predicted by RefSeq Genes 109.20200815 v1, NCBI. Non-Finnish European allele frequencies of UK Biobank are reported. The reference for each variant corresponds to the literature in which it was reported. Het; heterozygous, hom; homozygous, Geno; genotype detected in this population, LoF; loss of function, AGS; Aicardi-Goutières syndrome, CGD; Chronic Granulomatous disease, CVID; Common Variable Immunodeficiency, EDS; Ehlers Danlos syndrome, N.; number of individuals, SAPHO; synovitis, acne, pustulosis, hyperostosis and osteitis, SLE; Systemic lupus erythematosus, reported geno; the genotype reported in the corresponding reference, VUS; variant of unknown significance, VMLDS; Van Maldegem syndrome. (1) Hargreaves et al 2021, (2) Infante et al 2022, (3) Heddar et al., 2022, (4) Alaqeel et al, 2019, (5) Ferrari et al., 2023, (6) Atschekzei et al., 2022, (7) Offer et al., 2010, (8) Koehler et al., 2007, (9) Kiyozumi et al., 2019, (10) Van De Geer et al., 2018, (11) Walker et al., 2011, (12) Rice et al., 2007, (13) Weerakkody et al., 2016, (14) Pulvirenti et al., 2016, (15) Dziurdzik et al., 2020. (38,46,53,55,56,63–72).

### Variants within Drug Response or Toxicity Genes

A total of four variants within genes *ABCC2, DPYD, JAK1* and *NCF4* across four individuals were detected within genes that were reported on ClinPGX as related to response or toxicity for the drugs abatacept, adalimumab, anakinra, canakinumab, etanercept, golimumab, infliximab, methotrexate, rituximab, tocilizumab. These four variants have been previous reported in the literature, Table 5. Additionally, there are 35 novel variants within 29 genes from the drug ClinPGX search that are presented in Table 5. In total variants from the drug response or toxicity gene list were detected in 32 individuals.

**Table 5.**
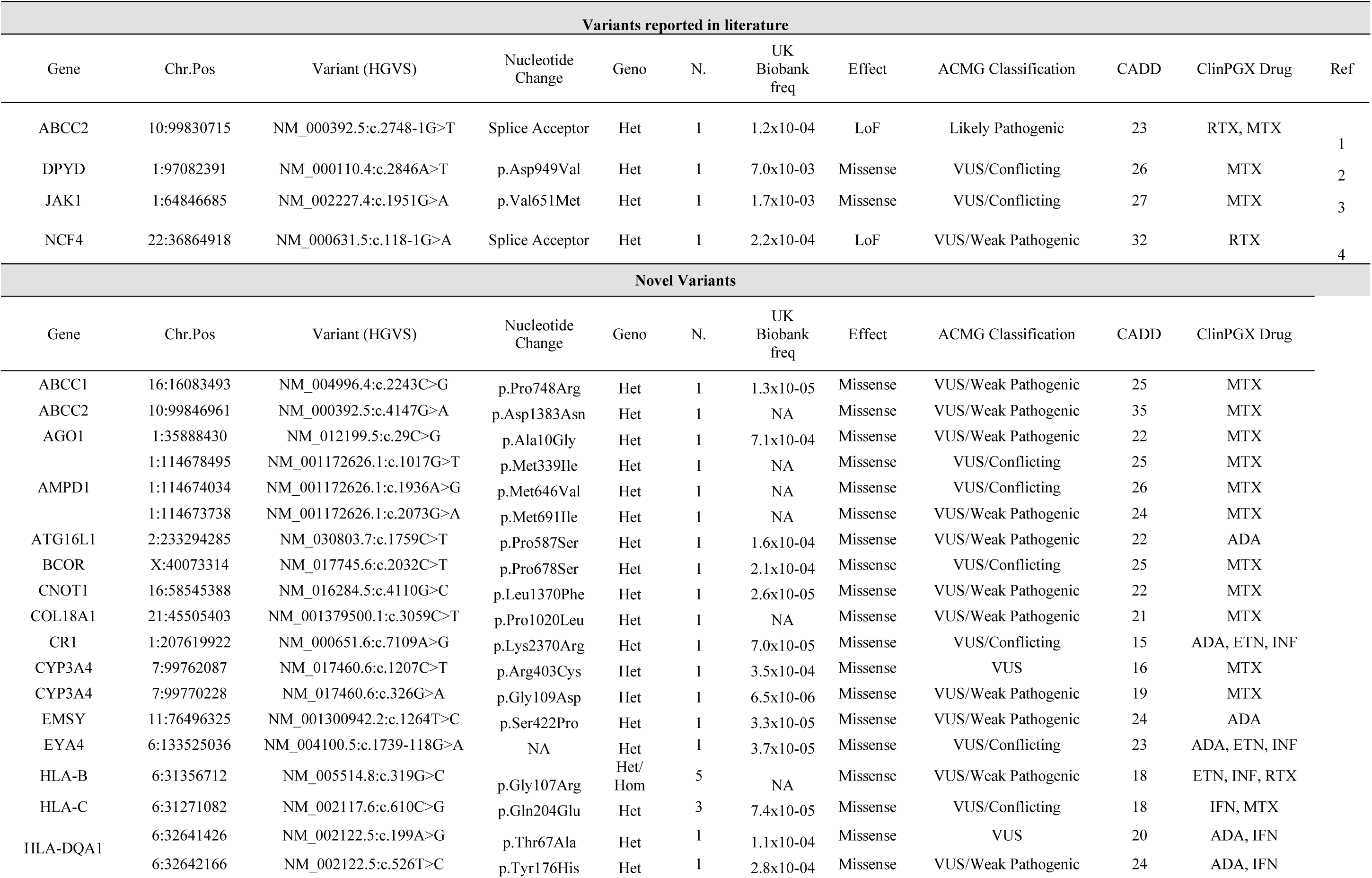
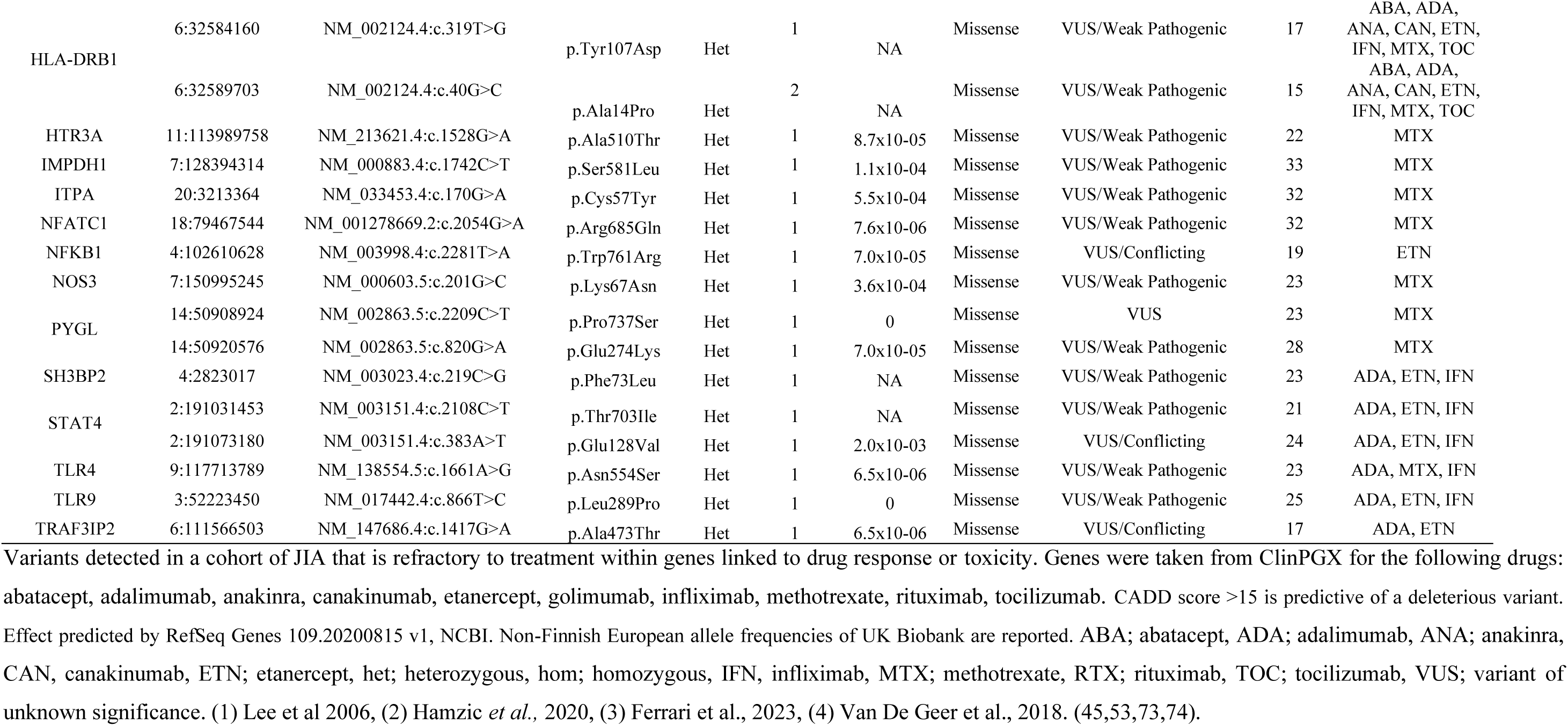
Variants within genes linked to toxicity or response of JIA drugs. Variants detected in a cohort of JIA that is refractory to treatment within genes linked to drug response or toxicity. Genes were taken from ClinPGX for the following drugs: abatacept, adalimumab, anakinra, canakinumab, etanercept, golimumab, infliximab, methotrexate, rituximab, tocilizumab. CADD score >15 is predictive of a deleterious variant. Effect predicted by RefSeq Genes 109.20200815 v1, NCBI. Non-Finnish European allele frequencies of UK Biobank are reported. ABA; abatacept, ADA; adalimumab, ANA; anakinra, CAN, canakinumab, ETN; etanercept, het; heterozygous, hom; homozygous, IFN, infliximab, MTX; methotrexate, RTX; rituximab, TOC; tocilizumab, VUS; variant of unknown significance. (1) Lee et al 2006, (2) Hamzic *et al.,* 2020, (3) Ferrari et al., 2023, (4) Van De Geer et al., 2018. (45,53,73,74).

## Discussion

To our knowledge, this is the first in-depth investigation of genetic variation in a population of JIA with refractory disease. We have detected evidence that this population carries genetic risk variants for paediatric-onset diseases that can present with symptoms similar to JIA as well as variants for risk for rare forms of JIA and candidate variants for drug response or toxicity to JIA therapies. It is important to note that due to the historical samples used in this study, longitudinal data were not available and updated information on diagnoses and treatment response could not be collected.

One variant was detected in the same genotype previously reported in the literature; p.Arg999His of *RYR1* is pathogenic in a heterozygous genotype for Multi-Minicore disease (35). *RYR1* encodes a skeletal muscle ryanodine receptor. Previously, the p.Arg999His variant was reported in two heterozygous siblings with Multi-Minicore disease and central core disease. Multi-Minicore disease is a rare muscle condition that causes muscle weakness and wasting, which primarily affects skeletal muscle (35) The carrier of p.Arg999His would need to undergo further diagnostic testing for Multi-Minicore and central core disease. Two variants detected within *RNASEH2B* were previously detected for SLE and Aicardi-Goutières syndrome (AGS), however, the genotype for either was not reported in the literature. Variant p.Asp105Ala within *RNASEH2B* was reported to have a minor allele frequency (MAF) of 0.08 in a cohort of SLE (36). SLE is an autoimmune disorder, which can include symptoms such as fever, malaise, arthralgias, non-erosive arthritis, myalgias, headache and fatigue (36,37). Additionally, loss of function (LoF) variant p.Ser28Ter of *RNASEH2B* is reported in a study of Aicardi-Goutières syndrome (AGS) (38). AGS is an interferonopathy with autoinflammatory features (39). Patients with AGS can have joint involvement, including lupus-like non-erosive arthritis or progressive arthropathy with joint contractures along with fever and systemic inflammation (40,41).

Nine further variants were detected in this study in genes for other paediatric diseases, however, not in the same genotype reported in the literature (Table 2). One example is variants p.Ile1148Thr and p.Gln1142His within *ATP7B,* which were detected in one individual in this study and are pathogenic for Wilson’s disease when in combination with a third variant, p.Met769Val (32). Wilson’s disease is a rare disorder of copper metabolism, which causes the toxic accumulation of copper in tissues in the liver, brain and other tissues leading to toxic effects (42). Variant p.Met769Val was not detected in the individual in this study, however there were an additional six *ATP7B* variants detected that did not pass the filtering pipeline. While musculoskeletal symptoms are unusual in Wilson’s disease, one case study reported a case of oligoarticular JIA not responding to standard treatment that was later diagnosed as Wilson’s disease. It is theorised that recurrent microtrauma and copper toxicity in the joints resulted in JIA-like joint symptoms (42). Further detailed information of each variant in can be found in the Supplementary material.

A total of 17 rare variants were detected within genes linked to arthritis phenotypes using OMIM phenotype match in VarSeq. While Table 3 presents rare variants within genes typical of JIA and autoimmune disease such as *HLA-B* and *HLA-DRB1*, there are some interesting novel variants for monogenic forms of JIA. Of particular interest is the one patient carrying two novel *LRBA* variants. *LRBA,* lipopolysaccharide-responsive and beige-like anchor protein, is a gene known to be causal for a LRBA deficiency, an immune deficiency in which familial case studies report JIA as a phenotype of the disease (43). In particular, the phenotype can be early-onset, severe and chronic (44).

A total of four variants within the genes *ABCC2, DYPD, JAK1* and *NCF4* linked to drug response or toxicity on ClinPGX were also detected in this cohort. ClinPGX had listed these genes in response or toxicity to methotrexate or rituximab. However, there have been no publications that specifically report the link of these variants to methotrexate or rituximab. Interestingly, splice variant c.2748-1G>T of *ABCC2* is reported to be pathogenic for early onset of Dubin-Johnson syndrome, a rare autosomal recessive disease of chronic or intermittent conjugated hyperbilirubinemia. The study reported that the variant is predicted to truncate the multidrug-resistance-associated protein 2 gene product and evidenced the absence of protein expression through immunohistochemical staining (45). *JAK1* variant p.Val651Met was detected in two “relapse” samples in a study of refractory Philadelphia negative acute lymphoblastic leukaemia, where the authors suggested the variant as a driver of relapse (46).

This study has utilised WES to detect rare variants in loci which may underlie monogenic diseases or drug response in a population of JIA patients with refractory disease. Using this method, we were able to detect rare and likely pathogenic variants in exons in a cohort of JIA that would not have been detected using genetic association methods due to small sample sizes and rarity of the variants. However, it is important to recognise the limitations of this study. One limitation of this study is the way in which refractory JIA was defined. In defining refractory disease in JIA, we aimed to identify cases of refractory inflammatory disease. EULAR has recently defined “difficult to treat” RA, which forms the basis for future research in adult rheumatology (47) however, such a definition is yet to be developed in paediatric rheumatology. Due to this, a definition for refractory JIA had to be devised for this study. This definition therefore included multiple use of biologic drugs which may have stopped for other reasons beyond uncontrolled disease. Consequently, some cases may not have true refractory inflammatory disease. We also applied this definition across all ILAR subtypes. JIA is a heterogeneous disease with seven ILAR subtypes, therefore treatment options to achieve low disease activity can differ. Consequently, it could be that refractory disease should be defined differently between JIA subtypes. Another limitation to this study is that there was not a healthy control population for a comparison study. The authors of the study attempted to utilise RV-EXCALIBER to conduct an aggregated rare variant association study using publicly available gnomAD WES data (48). However, the study was underpowered due to small sample size. Therefore, the study would benefit from a control WES population to validate the results. Finally, the ethic approvals of the data collections and analyses in the UK JIA cohorts do not allow for individual data to be fed back to physicians or patients themselves. This meant that further diagnostic tests, sequencing or protein function tests could not be completed, and further clinical details of the patient could not be obtained to validate our results. Therefore, it is important to note that the clinical significance of findings is not known at this stage.

Genetic testing of patients is a rapidly evolving field, with genetic diagnoses offering a means to advise treatment options and greater access to therapies. The importance of sequencing for progression in medicine is evidenced in the recent *Fit For The Future* initiative where there are plans to sequence the genomes of 100,000 newborn babies in the UK, which aims to drive healthcare reform (49). Sequencing could aid in finding a biomarker for diagnosis or treatment response for JIA, where diagnosis is still dependent on clinical assessment of symptoms (50). This study presents an exploration of rare variants in a cohort of JIA that is refractory. Early evidence of genetic variants for other paediatric diseases was detected in a small subset of the population studied. One variant was detected in a pathogenic genotype as previously reported in literature, however, further clinical data is not available for this patient. The authors also report a patient that carries two *LRBA* variants, which could suggest a rare subtype of familial JIA. Additionally, the study detected 39 variants in drug response or toxicity genes reported on ClinPGX for JIA therapies. The findings of this study would have been more readily interpreted had a comparator group of healthy controls been available. Therefore, further research is required to understand the presence of such variants in JIA and refractory disease. However, this study highlights that sequencing of refractory JIA cases in the future is important to aid diagnoses. Given the increasing availability, sequencing patients with JIA that is refractory could be paramount in understanding disease that does not response to treatment whether that be due to complex disease or other diagnostic possibilities. However, the challenge of interpreting the role of variants detected and their functional relevance remains.

## Data Availability

All data produced in the present study are available upon reasonable request to the authors.

## Acknowledgments and affiliations

CLUSTER is supported by grants from the Medical Research Council (MRC) [MR/R013926/1] and Versus Arthritis [Grant: 22084], Great Ormond Street Hospital Children’s Charity [VS0518], and Olivia’s Vision. This work is supported by the NIHR GOSH BRC, the NIHR Manchester Biomedical Research Centre, the NIHR GOSH Biomedical Research Centre and the British Society for Rheumatology (BSR), and the “UK’s Experimental Arthritis Treatment Centre for Children, supported by Versus Arthritis (grant: 20621)”. The views expressed are those of the author(s) and not necessarily those of the NHS, the NIHR or the Department of Health. Wedderburn is additionally supported by Versus Arthritis (grant: 21593) at the Centre for Adolescent Rheumatology Versus Arthritis. Hyrich is additionally supported by the Centre for Epidemiology Versus Arthritis (grant: 21755) and the Centre for Genetics and Genomics Versus Arthritis (grant: 21754) at the University of Manchester, UK. LKF is supported by Arthritis UK (23126).

This study acknowledges the use of the following UK JIA cohort collections: The Biologics for Children with Rheumatic Diseases (BCRD) study (funded by Arthritis Research UK grant: 20747); The British Society for Paediatric and Adolescent Rheumatology Etanercept Cohort Study (BSPAR-ETN) (funded by a research grant from the British Society for Rheumatology (BSR); BSR has previously also received restricted income from Pfizer to fund this project; Childhood Arthritis Prospective Study (CAPS) (funded by Versus Arthritis UK, grant: 20542); Childhood Arthritis Response to Medication Study (CHARMS) (funded by Sparks UK, reference 08ICH09; and the Medical Research Council, reference MR/M004600/1), United Kingdom Juvenile Idiopathic Arthritis Genetics Consortium (UKJIAGC). This study also acknowledges the use of the following two UK-wide JIA-associated uveitis clinical trials: the SYCAMORE Trial (funded by Arthritis Research UK, grant: 19612 and the National Institute of Health Research Health Technology Assessment, grant: 09/51/01); and the APTITUDE Trial (funded by Arthritis Research UK, grant: 20659).

## Members of the CLUSTER Consortium are as follows

Prof Lucy R. Wedderburn, Ms Zoe Wanstall, Ms Vasiliki Alexiou, Mr Fatjon Dekaj, Ms Bethany R Jebson, Dr Melissa Kartawinata, Ms Aline Kimonyo, Ms Eileen Hahn, Ms Genevieve Gottschalk, Ms Freya Luling Feilding, Ms Alyssia McNeece, Ms Fatema Merali, Ms Elizabeth Ralph, Ms Emily Robinson, Ms Emma Sumner (UCL GOS Institute of Child Health, London); Prof Andrew Dick, (UCL Institute of Ophthalmology, London); Prof Michael W. Beresford, Dr Emil Carlsson, Dr Joanna Fairlie, Dr Jenna F. Gritzfeld, Dr Oliver McClurg, Dr Karen Rafferty (University of Liverpool); Prof Athimalaipet V Ramanan, Ms Teresa Duerr (University Hospitals Bristol and Weston NHS Foundation Trust); Prof Michael Barnes, Ms Sandra Ng, (Queen Mary University, London); Prof Kimme Hyrich, Prof Stephen Eyre, Prof Soumya Raychaudhuri, Prof Wendy Thomson, Dr John Bowes, Ms Jeronee Jennycloss, Ms Saskia Lawson-Tovey, Dr Paul Martin, Prof Andrew Morris, Dr Stephanie Shoop-Worrall, Dr Samantha Smith, Dr Michael Stadler, Dr Damian Tarasek, Dr Melissa Tordoff, Dr Annie Yarwood (University of Manchester); Dr Chris Wallace, Dr Wei-Yu Lin (University of Cambridge); Prof Nophar Geifman (University of Surrey); Dr Sarah Clarke (School of Population Health sciences and MRC Integrative Epidemiology Unit, University of Bristol). Dr Thierry Sornasse (AbbVie Inc.) Dr Robert J Benschop, Dr Rona Wang (Eli Lilly) Daniela Dastros-Pitei MD, PhD, Sumanta Mukherjee, PhD (GlaxoSmithKline Research and Development Limited.) Dr Michael McLean, Dr Anna Barkaway (Pfizer) Dr Peyman Adjamian (Swedish Orphan Biovitrum AB (publ) (Sobi)) Helen Neale (UCB Biopharma SRL.) The CLUSTER Champions.

We are grateful to the CLUSTER champions and members of the CLUSTER consortium, who have provided valuable feedback for this work and for the future dissemination of this work to patients and parents. The authors would like to acknowledge the assistance given by Research IT and the use of the Computational Shared Facility at The University of Manchester. Ethical approvals were gained from the Northwest Greater Manchester Central Research Ethics Committee (BCRD), West Midlands Multicentre Research Ethics Committee (BSPAR-ETN), Northwest Multicentre Ethics Committee (CAPS: REC/02/8/104, IRAS 184042) and the Bloomsbury/Central London Research Ethics Committee (CHARMS: REC 05/Q0508/95, IRAS 172219). No additional ethical permissions were required for this analysis. Written informed consent was provided by guardians of participants and age-appropriate consent/assent was provided by participants themselves, where appropriate. All authors have completed the ICMJE uniform disclosure at http://www.icmje.org/disclosure-of-interest/ and declare: MT has received research grants from SOBI and AbbVie, AVR has received consulting fees and honorariums from AbbVie, Eli Lilly, UCB, Astra Zeneca, Pfizer, Roche, Novartis, Alimera and SOBI, KH has received grants from Pfizer and BMS unrelated to work in JIA and honorariums from AbbVie and is an Associate Editor at ARD, LRW has received research grants from MRC, Versus Arthritis, AbbVie, SOBI, UCB, GSK and Great Ormond Street Children’s Charity and honorariums from Pfizer and Advanced Targeted Therapies.

We are grateful to the CLUSTER champions and members of the CLUSTER consortium, who have provided valuable feedback for this work and for the future dissemination of this work to patients and parents. The authors would like to acknowledge the sample processing team at The University of Manchester Centre for Genetics and Genomics Versus Arthritis; Ruairi McErlean.

## Notes

### Competing Interest Statement

LRW would like to declare the following UK Medical Research Council MR/R013926/1, Versus Arthritis Charity, Great Ormond Street Children's Charity VS0518, In kind contributions to CLUSTER AbbVie, UCB, Pfizer GSK and SOBI, AbbVie research grant and SOBI research grant to UCL and academic partners. NIHR Great Ormond Street Biomedical Research Centre to UCL. Advanced Targeted therapies meeting speaker honorarium 2022, Speaker fees Pfizer 2023, Speaker feed Pfizer 2024 to UCL. MT would like to declare The CLUSTER Consortium industry partner funding from SOBI and AbbVie awarded to the University of Manchester. SE would like to declare a grant from Janssen for unrelated work awarded to the University of Manchester. AR would like to declare consulting fees from Abbvie, Eli Lilly, UCB and Astra Zeneca and payment for honoraria from Abbvie, Eli Lilly, Pfizer, Roche, Novartis, UCB, Alimera and SOBI. KH would like to declare Pfizer and BMS grants paid to University of Manchester for research unrelated to JIA, fees for speaking at educational meetings from Abbvie to the University of Manchester. KH has a role as associate Editor at ARD.

### Author Declarations

Ethical approvals were gained from the Northwest Greater Manchester Central Research Ethics Committee (BCRD), West Midlands Multicentre Research Ethics Committee (BSPAR-ETN), Northwest Multicentre Ethics Committee (CAPS: REC/02/8/104, IRAS 184042) and the Bloomsbury/Central London Research Ethics Committee (CHARMS: REC 05/Q0508/95, IRAS 172219). No additional ethical permissions were required for this analysis. Written informed consent was provided by guardians of participants and age-appropriate consent/assent was provided by participants themselves, where appropriate.

